# 24-hour movement behaviors and cardiometabolic health in adults with type 2 diabetes: a comparative cross-sectional and longitudinal analysis

**DOI:** 10.1101/2025.11.14.25340217

**Authors:** Lotte Bogaert, Marieke De Craemer, Eveline Dirinck, Patrick Calders, Bruno Lapauw, Iris Willems

**Affiliations:** Ghent University, Department of Rehabilitation Sciences, Ghent, Belgium; Research Foundation Flanders, Brussels, Belgium; Department of Endocrinology, Antwerp University Hospital & University of Antwerp, Antwerp, Belgium; Department of Internal Medicine and Pediatrics & Department of Endocrinology, Ghent University & Ghent University Hospital, Ghent, Belgium

## Abstract

**Introduction:** Meeting the recommended guidelines for physical activity (PA), sedentary behavior (SB) and sleep, collectively referred to as 24-hour movement behaviors (24h-MBs), is crucial for type 2 diabetes mellitus (T2DM) management and is associated with favourable health outcomes. However, it is suggested that adults with T2DM spend more time in SB and less time in PA compared to adults without diabetes.

**Objectives:** This study aims to compare 24h-MBs between adults with and without T2DM (i.e. controls with similar characteristics except for having T2DM), investigate how this is associated with cardiometabolic health, and assess changes in 24h-MBs after two years of follow-up (FU) in adults with T2DM.

**Design:** Cross-sectional and longitudinal study

**Setting:** Community-dwelling adults with T2DM and controls in Belgium.

**Primary outcome measures:** This study took place between September 2021 and December 2023. The 24hMBs were measured using accelerometers (Actigraph wGT3X+); cardiometabolic variables (adiposity, blood pressure and advanced glycation endproducts) were collected in both groups. In adults with T2DM, fasting blood samples were collected at baseline and second FU. Compositional data analysis was used to explore group differences in 24h-MBs using MANOVA, and regression models analysed associations with cardiometabolic health. Changes in 24h-MBs over time in adults with T2DM were assessed using a linear mixed model.

**Results:** Fifty-two adults with T2DM (mean age 63.2 standard deviation (SD) 10.6) and 74 controls (mean age 62.7SD 9.4) were included in the cross-sectional analysis. The 24h-MBs of adults with T2DM differed significantly from the controls (p=0.026). Adults with T2DM spent significantly less time in light (-34.7 min/day) and moderate-to-vigorous PA (MVPA) (-24.1 min/day) compared to controls. In adults with T2DM, reallocating 30 minutes from any behavior to MVPA was associated with a significant increase in HDL-cholesterol (sleep: 5.05 mg/dl [2.45; 7.80], ES=0.53; SB: 4.53 mg/dl [1.93; 7.27], ES=0.47; LPA: 5.29 mg/dl [2.07; 8.73], ES=0.55) whereas in the control group significant decreases in waist circumference where found when reallocating 30 minutes from SB to sleep (2.42 cm [0.86-3.97], ES=0.34). Thirty-seven (mean age 65.0 SD 9.5) and 22 (mean age 67.0 SD 7.7) adults with T2DM provided valid data after one and two years of follow up, respectively. No significant changes in 24h-MBs were found after one (p=0.93) or two-year (p=0.79) FU among adults with T2DM .

**Conclusion:** Adults with T2DM have a less favorable 24h-MB composition compared to adults without T2DM indicating the need for additional effort to achieve and maintain the guidelines. Despite the limited associations found, time reallocations from other behaviors to MVPA theoretically suggest the biggest health benefits.

**Trial registration:** NCT04993482 (ClinicalTrials.gov)

**Strengths and limitations of this study:**

- This study uses objectively collected sleep, sedentary behavior and physical activity data, which limited the risk of recall- and social desirability bias.
- This study includes longitudinal data which provides valuable insights into behavioral changes over time.
- Participants could choose whether the data collection took place at the Ghent University Hospital or at the participant’s home. The location could potentially influence certain measurements, such as BP.
- Since no blood samples were taken in the control group, it is possible that these individuals have prediabetes.

## Background

Type 2 diabetes mellitus (T2DM) is a global pandemic, affecting over 500 million people worldwide in 2021. Projections suggest that the prevalence of T2DM is going to double by 2050 [1]. This chronic disease is characterized by a significant risk for complications, with cardiovascular disease (CVD) as the leading cause of comorbidity and death in adults with T2DM [1, 2]. To delay or prevent the onset of CVD, an effective management of T2DM is crucial.

An important component of T2DM management is adhering to the recommended guidelines for physical activity (PA) (i.e. at least 150 minutes moderate to vigorous PA/week), limiting sedentary behavior (SB) (i.e. less than 8 hours/day), and obtaining sufficient sleep (i.e. 7-9 hours/night) [3, 4]. These behaviors have been shown to support T2DM management (e.g., glucose control) and to affect cardiovascular risk factors (e.g., blood pressure (BP), lipid profile) [3, 5–9]. Traditionally, PA, SB and sleep have been investigated in isolation with a great focus on moderate-to-vigorous PA (MVPA) [10]. However, PA, SB and sleep occur within a 24-hour day, making them time- and interdependent and are therefore referred to as 24-hour movement behaviors (24h-MBs). An integrated approach considering the interdependence of these behaviors should be used to formulate health-promoting strategies for people with T2DM, which is also highlighted in the recent Standards of Care in Diabetes [4, 11].

Recent studies have examined this 24h-MBs paradigm in adults with T2DM [12–14]. One study involving over 1000 adults with T2DM found significant differences in 24h-MBs based on weight status with adults classified as obese showing more SB and less PA than overweight or normal weight adults [15]. Furthermore, the 24h-MB composition of adults with T2DM is associated with various health outcomes. More time spent in MVPA relative to the other behaviors correlates with an improved body composition (i.e. body mass index (BMI), waist-to-hip ratio (WHR), waist circumference (WC)) and cardiometabolic health (HDL-cholesterol, triglycerides, insulin resistance (IR)). In addition, higher proportions of light PA (LPA) are associated with lower triglyceride and plasma insulin levels, while more time spent in SB is associated with worser lipid profiles [14–16]. The effects of sleep duration are mixed: reallocating sleep time into more MVPA is shown to be beneficial for health outcomes only among long sleepers (9.3-h/night), while reallocating SB to sleep in short- to average sleepers (7.7-h/night) is associated with reductions in BMI and WC [15].

Until now, no studies compared 24h-MB compositions between adults with T2DM and adults without diabetes. It is hypothesized that these behaviors will differ based on previous isolated behavior studies showing that adults with T2DM tend to allocate less time to PA, more time to SB and experience a less optimal sleep compared to their adults without diabetes [17–20]. Moreover, it is not yet explored if there are differences in associations with cardiometabolic health between these groups. Investigating (possible) differentiation between these groups could provide valuable insights into whether extra efforts regarding 24h-MB recommendations are needed for adults with T2DM. Last, little is known about the possible long-term changes in 24h-MBs among adults with T2DM. Understanding the stability of 24h-MBs in this population could provide important information for future interventions.

Based on current knowledge gaps, this study aims to compare 24h-MBs of adults with and without T2DM (aim 1), explore cross-sectional associations between 24h-MBs and cardiometabolic variables for both groups (aim 2) and describe the annual change or stability in 24h-MB compositions over a 2-year period among adults with T2DM (aim 3).

## Method

### Design

Both cross-sectional and longitudinal data were collected in Flanders, Belgium from September 2021 to December 2023. The recruitment of adults with T2DM and the control group for the cross-sectional study began in September 2021 and ended in December 2023. Following the first assessment, adults with T2DM included in the cross-sectional study in 2021 or 2022 were invited to participate in longitudinal study involving up to two annual measurements (from September 2022 until December 2023). The control group was not invited for the longitudinal study. The longitudinal study was completed by December 2023.

### Participants

Adults with T2DM were eligible if they were ≥18 years, were diagnosed with T2DM by a physician or had an HbA1c >6.5%, and had no physical limitations (e.g. amputation), cognitive disabilities (e.g. dementia), or other conditions that would hinder normal PA. Recruitment of adults with T2DM occurred through Ghent University Hospital, the Diabetes Liga, and the distribution of flyers in public places (e.g. general practitioners’ waiting rooms). Controls, who met the same inclusion criteria but did not have a diagnosis of T2DM (verbally confirmed to have no known T2DM diagnosis), were recruited through flyers in public places and convenience sampling within researchers’ networks.

This study was approved by the local Ethics Committee (Ghent University Hospital, Belgium [BC-10189]), and written informed consent was obtained from all participants.

### Patient and Public involvement statement

Participants were not involved in the development of this study.

### Data collection

Data were collected either at Ghent University Hospital or at the participant’s home. During the visit, several cardiometabolic variables were measured and participants were instructed on how to wear the accelerometer, complete the sleep diary, and fill out the online questionnaires. After a wear period of seven days, the accelerometer and the sleep diary were returned via a prepaid postal package. The same measurements were taken during the follow-up assessments of the longitudinal study in adults with T2DM.

### Participant characteristics

To provide a description of our population, information regarding sex, age, household income, work status, education (up to secondary vs bachelors or above), smoking status, medication intake and diabetes duration (only in adults with T2DM) was collected through an online questionnaire. Medication was classified as: lipid-lowering medication, BP lowering medication, (number of) glucose lowering medication, anticoagulant medication, insulin or other. Based on previous research, some variables (i.e., sex, age, education, smoking, income, and environment) were also included as covariates in both the cross-sectional (aim 1 and 2) and longitudinal analysis (aim 3) [21, 22].

### 24-hour movement behaviors features

Data on 24h-MBs were objectively collected using an accelerometer (ActiGraph wGT3X+BT) for seven consecutive days. During waking hours, the accelerometer was worn on the right hip. At night the device was switched to the non-dominant wrist. The accelerometer was removed for water-based activities. To validate sleep time and (non)wear activities, the accelerometer was supplemented with a written diary to record bed- and wake-up times. The information of this diary was used in later data-analysis to obtain the most valid measurement of sleep and SB (i.e. if non-wear time was not defined, it would be incorrectly added to SB). The accelerometer was initialized via Actilife v6.13.4 software with a sampling frequency of 30 Hz and an epoch length of 60 seconds. Raw data were downloaded using Actilife and further processed in the R-package GGIR (version 3.0.2) to estimate our primary outcome, i.e. 24h-MBs [23]. GGIR uses an autocalibration algorithm that checks and corrects for calibration errors in triaxial accelerometer signals. Actigraph files (n=1) with post calibration errors greater than 0.01*g* were excluded [23]. To have a valid estimation of the 24h-MBs, non-wear time was detected using 15-minute time blocks based on the characteristics of the 60-minute time window. A block was defined as non-wear if at least two of the three accelerometer axes had a standard deviation of accelerations of 13.0 mg or less (1 milli gravity-based acceleration unit (*mg*) = 0.00981 m/second²) or if the value range of acceleration was less than 50 *mg* [24]. Days were considered as valid if there was ≥16 hours wear time, and participants with at least three valid weekdays and one valid weekend day were included in the analysis. Sleep duration was calculated using the sleep diary or the GGIR default Heuristic Algorithm looking at Distribution of Change in Z-angle (HDCZA) if there was no sleep diary available. The HDCZA algorithm categorizes periods with no change larger than 5 degrees in the non-dominant arm angle over at least 5 minutes as sleep [25]. Non-sleep time (i.e. total wear time – sleep time) was subdivided in activity categories (i.e. SB, LPA or MVPA) based on the Mean Amplitude Deviation (MAD) cut-points provided by Vähä-Ypyä et al: SB (>16.7 *mg*), LPA (16.7-90.9 *mg*), moderate PA (91-414 *mg*) and vigorous PA (≥414 *mg*) [26, 27]. MVPA was then computed as the sum of moderate and vigorous PA.

For each participant, the average number of minutes of daylight (sunrise to sunset) corresponding to the period of the accelerometer wear was calculated. This measure was included as a covariate to control for potential confounding effects in both the cross-sectional (aim 1 and 2) and longitudinal analysis (aim 3).

Additionally cut-point independent metrics such as average acceleration and intensity gradient were calculated by GGIR [28]. The average acceleration represents the overall activity volume represented in m*g* [28]. The intensity gradient refers to the intensity distribution over a 24h day. A smaller intensity gradient (more negative) reflects a more sedentary profile [28].

### Cardiometabolic variables

To address the second aim of this study, several cardiometabolic variables (primary outcomes) were measured. Height and weight were measured twice using a portable stadiometer (Seca 213, to the nearest 0.1 cm) and electronic scale (Tefal Premia type 5243, to the nearest 0.1 kg), respectively, with participants wearing light clothes without shoes. BMI was calculated by dividing the mean weight (in kg) by the square of the mean height (in meters). WC was measured twice at the mid-point between the highest point of the iliac crest and the lower rib, and hip circumference was measured twice around the widest portion of the buttocks. Both measurements were taken to the nearest 0.1 cm. WHR was computed by dividing the mean WC by the mean hip circumference. BP (in mm Hg) was measured twice, with an one-minute interval, with an automatic OMRON M6 Comfort device on the non-dominant arm after 10 minutes of rest. Skin Advanced Glycation End products (AGEs), expressed in arbitrary units (AU), were non-invasively measured via skin autofluorescence using an AGE reader (DiagnOptics BV) based on three consecutive measurements on the dominant arm of the participant. These measurements were taken twice with a one-minute interval.

A fasting blood sample was collected in adults with T2DM. Participants were instructed to abstain from eating and drinking (except water) for a minimum of ten hours. Lipid profile (total cholesterol (TC), high-density lipoprotein cholesterol (HDL-C), low-density lipoprotein cholesterol (LDL-C), and triglycerides (TG)), blood glucose control (HbA1c (in mmol/mol and in %) and fasting glucose levels) and serum insulin levels were determined. TC, HDL-C and TG were routinely determined using standard laboratory assays (COBAS 8000 modular analyser series, Roche Diagnostics, Mannheim, Germany). LDL-C was calculated using the Martin formula. Serum levels of fasting glucose were analysed by the hexokinase method (COBAS, Roche Diagnostics) and HbA1c analyses were performed on a Tosoh HLC-723 G8 high-performance liquid chromatography (HPLC) analyser. Insulin resistance (Homeostatic Model Assessment of Insulin Resistance (HOMA-IR) and β-cell function (HOMA of Bèta-cell function (HOMA-B)) were calculated based on insulin levels and fasting glucose levels using the Homeostatic Model Assessment (HOMA) calculator (version 2.2.3, https://www.dtu.ox.ac.uk/homacalculator/). Both indices are dimensionless and therefore have no units.

### Data analysis

All data were analysed with R software version 4.3.2. Participants’ characteristics were reported as mean ±SD for continuous data and proportions (in %) for categorical data. To test differences in participant characteristics between the adults with T2DM and the control group, independent sample t-tests (continuous data) and chi-square tests (categorical data) were performed.

Compostional Data Analysis (CoDA) (R-package compositions) was used to transform the four distinct behaviors (sleep, SB, LPA and MVPA) into four sets of three isometric log ratio’s (ILRs) [29]. The combination of three ILRs represent the total composition independent of which behavior is placed at the numerator of z1. The formula below shows one set of three ILRs with SB as numerator of z1 [30].

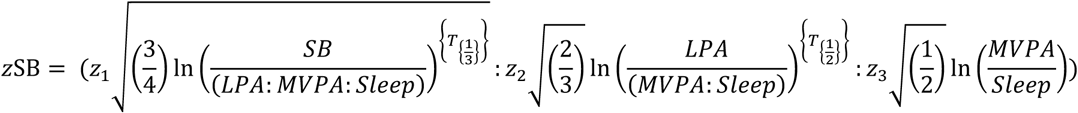

To compare the 24h-MB compositions between the two groups (aim 1), two MANOVA’s were performed with the ILRs’ coordinates as dependent variable and the grouping variable as independent variable. Both an unadjusted model (without confounders and covariate) and an adjusted model (corrected for sex, age, education, smoking, income, environment and daylight) were conducted. The adjusted model was used to assess and account for the potential confounding effects of these variables on the outcome.

For the second aim of this study, separate linear regression models were performed for each cardiometabolic health variable (i.e., BMI, WHR, AGEs, systolic BP, and diastolic BP in both groups; and TC, HDL-C, LDL-C, TG, HbA1c, fasting glucose, and insulin levels in adults with T2DM). In each model, the respective cardiometabolic health variable served as the dependent variable, and 24h-MB composition, expressed as ILR coordinates, was included as the independent variable. Assumptions were checked by evaluation of the linearity, homoscedasticity, interdependence of residuals and normality of residuals using the see and parameters packages [31, 32]. If one of the assumptions was not met, a log transformation was performed (AGE, diastolic blood pressure, BMI, WC (only in T2DM) and all blood parameters). A p-value correction was done using the Benjamini-Hochberg procedure to control for the multiple linear regression models and reduce type I errors. All regression models were adjusted for sex, age, education, smoking status, environment and daylight. If the cardiometabolic variable was significantly associated with the 24h-MB composition (p<0.05), theoretical time reallocation model predictions (one-v-one) were performed (codaredistlm package [33]). The average difference in the cardiometabolic variable was estimated when time (-30 to +30 min) in one behavior was replaced by time in another behavior while keeping the other behaviors constant. The absolute difference of change with respect to the mean were interpret. Log transformed cardiometabolic variables (i.e. HDL-cholesterol) were back transformed to interpret the results. An effect size was calculated for the significant associations by dividing the mean difference with the pooled standard deviation.

Regarding cut-point independent metrics (average acceleration and intensity gradient), linear regression models were built to explore the associations between cardiometabolic health (dependent variable) and average acceleration or intensity gradient (independent variable). Three models were built to explore the independent associations of both intensity and volume. Model 1 includes one of the two features (average acceleration or intensity gradient). Model 2 was additionally adjusted for sex, age, education, smoking status, environment and daylight. Model 3 was further adjusted for the alternate metric (average acceleration or intensity gradient depending on model 1) to test whether associations were independent of either feature.

To determine longitudinal changes in 24h-MB composition of adults with T2DM (aim 3), a linear mixed model was used with the ILRs as dependent variable presented in a long stacked format with dummy variables indicating the number of ILR (ILR1, ILR2, ILR3) as described by Lim and colleagues [34]. The independent variable in the model was time, represented by different timepoints (Timepoint one (T1) after one year of follow-up, timepoint two (T2) after two years). Random slopes were added at the log ratio level, grouped by participant ID and repeated within participant ID (random intercept) for each timepoint (lme4 package [35]) to account for repeated measurements over time, allowing each participant to have their own baseline ILR values (intercept) and individual patterns of change (slope) across the three timepoints. Fixed interaction effects between the timepoints (i.e. T1 and T2) and the dummy variable representing the ILR number were analysed to test for differences over time (MANOVA F-test). The model was adjusted for six confounders (i.e. sex, age, education, smoking, income, environment) and one covariate (i.e. daylight).

## Results

A total of 52 adults with T2DM and 74 control participants were included in the cross-sectional study (aims 1 and 2). Of these, 37 adults with T2DM took part in the longitudinal study at T1, and 22 adults with T2DM completed the longitudinal study at both timepoints (T1 and T2) (aim 3). Participants were lost to follow-up primarily due to withdrawal of consent, time constraints, or medical reasons (e.g. knee surgery). **Figure 1** presents a flow diagram illustrating participant recruitment and retention throughout the study.

**Figure 1.**
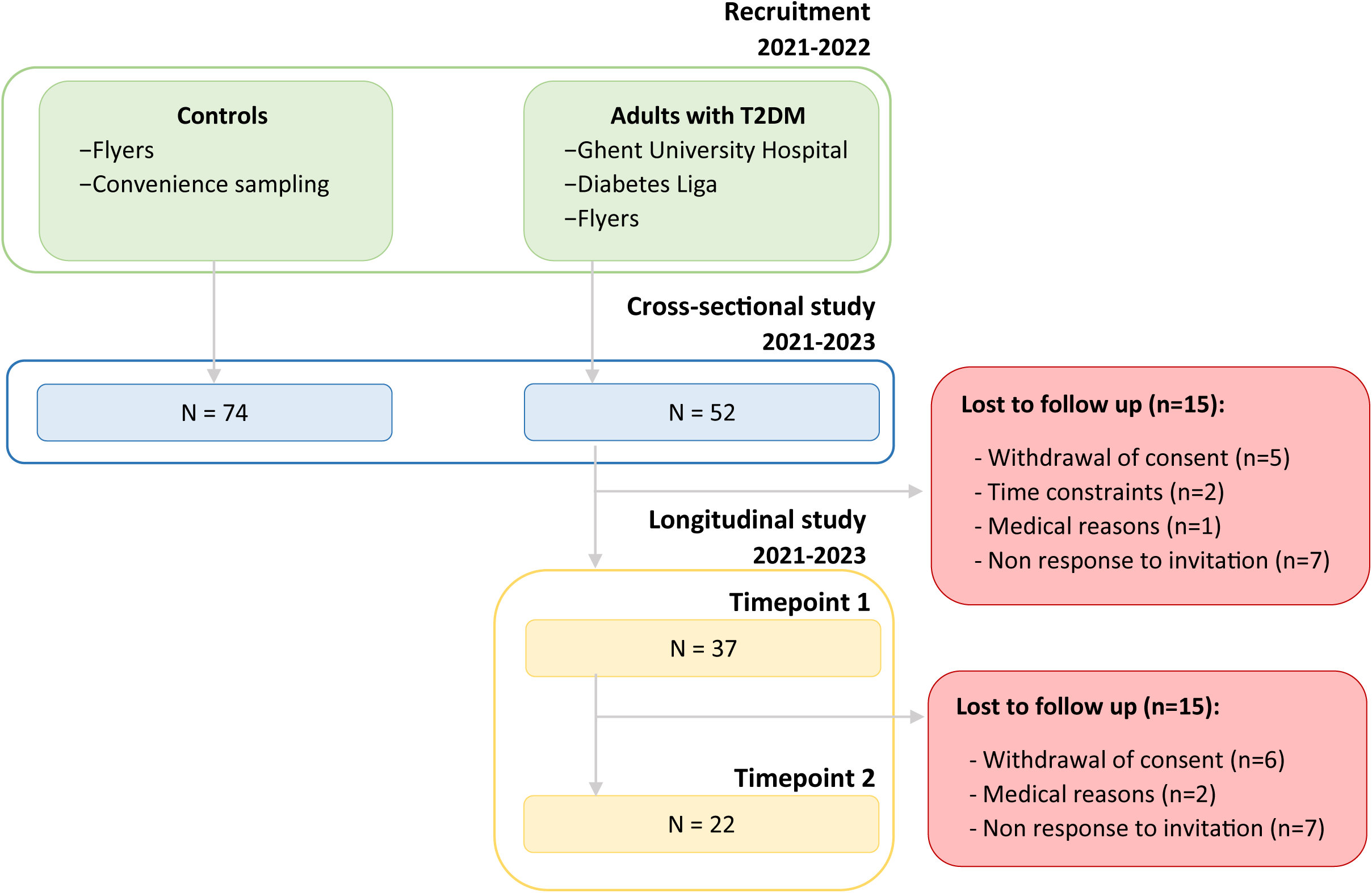
Flow diagram of recruitment and participant numbers in the cross-sectional and longitudinal study *Abbreviations*: T2DM: type 2 diabetes mellitus

### Participant characteristics

The mean age of the total sample was 62.9 (standard deviation (SD): 9.9) years with two thirds of the participants being male (68.3%). In adults with T2DM, the mean T2DM duration was 11.4 (SD: 6.7) years. Significant differences in body composition were observed between the groups: adults with T2DM had a significantly higher BMI (30.3 (SD: 5.4) versus 26.6 (SD: 4.1) kg/m²), WHR (1.0 (SD: 0.1) versus 0.9 (SD: 0.1)) and WC (107.4 (SD: 13.7) versus 96.6 (SD: 14.0) cm) compared to the controls (all p<0.001). Additionally, BP and skin AGEs differed significantly between the two groups with adults with T2DM having higher BP (systolic BP: 136.3 (SD: 17.8) versus 129.0 (SD: 14.8) mmHg), p=0.014; diastolic BP: 82.2 (SD: 11.1) versus 78.0 (SD: 13.7) mmHg, p=0.07) and more skin AGEs (2.7 (SD: 0.6) AU versus 2.5 (SD: 0.5) AU, p=0.021) compared to the controls. More detailed baseline characteristics are provided in **Table 1**.

**Table 1.** Study characteristics at baseline.

| Variable | Total sample (n=126) | T2DM (n=52) | Control (n=74) | P-value |
| --- | --- | --- | --- | --- |
| Age (in years) | 62.9 (SD 9.9) | 63.2 (SD 10.6) | 62.7 (SD 9.4) | 0.768 |
| Sex (n) |  |  |  | 0.998 |
| – Men | 86 (68.3%) | 36 (69.2%) | 50 (67.6%) |  |
| – Women | 40 (31.7%) | 16 (30.8%) | 24 (32.4%) |  |
| Environment (n) |  |  |  | 0.048* |
| – City | 38 (30.2%) | 21 (40.4%) | 17 (23.0%) |  |
| – Rural area | 87 (69.0%) | 30 (58.8%) | 57 (77.0%) |  |
| Education (n) |  |  |  | 0.004* |
| – Up to secondary | 50 (39.7%) | 28 (54.9%) | 22 (29.7%) |  |
| – Bachelors or above | 75 (59.5%) | 23 (44.2%) | 52 (70.2%) |  |
| Work status (n) |  |  |  | 0.149 |
| – Working | 55 (43.7%) | 18 (35.3%) | 37 (50.0%) |  |
| – Retired or unemployed | 70 (55.5%) | 33 (62.8%) | 37 (50.0%) |  |
| Household income (n) |  |  |  | 0.033* |
| – Low | 14 (11.1%) | 10 (19.6%) | 4 (5.6%) |  |
| – High | 109 (86.5%) | 41 (78.8%) | 68 (91.9%) |  |
| Smoking status (n) |  |  |  | 0.054 |
| – Ex-smoker | 31 (24.6%) | 17 (33.3%) | 14 (18.9%) |  |
| – Smoker | 6 ( 4.8%) | 4 (7.8%) | 2 (2.7%) |  |
| – Non-smoker | 88 (69.8%) | 30 (58.8%) | 58 (78.4%) |  |
| Diabetes duration (in years) |  | 11.4 (SD 6.7) | NA |  |
| Cardiometabolic health |  |  |  |  |
| BMI (in kg/m<sup>2</sup>) | 28.1 (±5.0) | 30.3 (±5.4) | 26.6 (±4.1) | <0.001* |
| BMI categories (n) |  |  |  | 0.002* |
| – Normal weight (>25kg/m<sup>2</sup>) | 38 (30.2%) | 9 (17.3%) | 29 (39.2%) |  |
| – Overweight (25-29.9 kg/m<sup>2</sup>) | 52 (41.3%) | 20 (38.5%) | 32 (43.2%) |  |
| – Obese (≥30 kg/m<sup>2</sup>) | 36 (28.6%) | 23 (44.2%) | 13 (17.6%) |  |
| Waist circumference (in cm) | 101.1 (SD 14.8) | 107.4 (SD 13.7) | 96.6 (SD 14.0) | <0.001* |
| Waist to hip ratio | 1.0 (SD 0.1) | 1.0 (SD 0.1) | 0.9 (SD 0.1) | 0.001* |
| Systolic blood pressure (in mm/Hg) | 132.0 (SD 16.4) | 136.3 (SD 17.8) | 129 (SD 14.8) | 0.014* |
| Diastolic blood pressure (in mm/Hg) | 79.8 (SD 12.8) | 82.2 (SD 11.1) | 78.0 (SD 13.7) | 0.07* |
| Advanced Glycation Endproducts (AU) | 2.6 (SD 0.5) | 2.7 (SD 0.6) | 2.5 (SD 0.5) | 0.021* |
| HbA1c (in mmol/mol) |  | 51.7 (SD 8.9) | NA |  |
| Insulin (in mU/l) |  | 17.5 (SD 14.4) | NA |  |
| Fasting glucose (in mg/dl) |  | 128.7 (SD 31.6) | NA |  |
| Total cholesterol (in mg/dl) |  | 151.5 (SD 39.1) | NA |  |
| HDL-cholesterol (in mg/dl) |  | 47.4 (SD 12.3) | NA |  |
| LDL-cholesterol (in mg/dl) |  | 83.5 (SD 29.6) | NA |  |
| Triglycerides (in mg/dl) |  | 125.9 (SD 75.5) | NA |  |
| HOMA-IR |  | 2.2 (SD 1.5) | NA |  |
| HOMA-B |  | 84.2 (SD 44.1) | NA |  |
| Medication intake |  |  |  |  |
| Medication (n yes) | 102 (80.9%) | 52 (100%) | 48 (64.7%) | <0.001* |
| Number of glucose lowering medication (n) |  |  |  |  |
| – <2 |  | 37 (71.1%) |  |  |
| – ≥2 |  | 15 (28.8%) | NA |  |
| Administering insulin (n yes) |  | 12 (23.1%) | NA |  |
| GLP1-receptor agonist medication (n yes) |  | 10 (19.2%) | NA |  |
| Lipid lowering medication (n yes) | 61 (48.4%) | 20 (39.2%) | 45 (62.5%) | 0.018* |
| Blood pressure lowering medication (n yes) | 58 (46.0%) | 18 (35.3%) | 50 (69.4%) | <0.001* |
| Anticoagulant medication (n yes) | 33 (26.2%) | 30 (58.8%) | 63 (87.5%) | 0.001* |
| Other medication (n yes) | 52 (41.3%) | 30 (57.7%) | 22 (29.7%) | 0.003* |
| Mean daylight (in minutes) | 591.4<br>(SD 113.3) | 509.8<br>(SD 52.1) | 648.8<br>(SD 109.5) | <0.001* |
| **24-hour movement behavior composition** |  |  |  |  |
| Sleep | 35.1 | 35.1 | 35.0 |  |
| Compositional mean in %<br>(min/day) | (505.2) | (505.6) | (503.5) | NA |
| Sedentary behavior | 43.9 | 46.0 | 42.3 |  |
| Compositional mean in %<br>(min/day) | (631.7) | (662.0) | (609.7) | NA |
| Light physical activity | 18.0 | 16.6 | 18.9 |  |
| Compositional mean in %<br>(min/day) | (258.5) | (238.6) | (272.8) | NA |
| Moderate to vigorous<br>physical activity | 3.1 | 2.3 | 3.8 |  |
| Compositional mean in %<br>(min/day) | (44.5) | (33.7) | (54.1) | NA |
| Average acceleration<br>(in mg) | 18.1 (SD 6.6) | 15.8 (5.4) | 19.7 (7.0) | 0.001* |
| Intensity gradient | -2.5 (SD 0.3) | -2.52 (SD 0.3) | -2.45 (SD<br>0.3) | 0.144 |

\*p<0.05, continuous variables were represented as mean (standard deviation (SD)), categorical variables as number (percentage). Information on missing data is provided in supplementary file 1.
*Abbreviations:* T2D: type 2 diabetes; SD: standard deviation; BMI: body mass index; LDL-cholesterol: low-density lipoprotein cholesterol; HDL-cholesterol: high-density lipoprotein cholesterol; HOMA-IR: Homeostatic Model Assessment of Insulin Resistance; Homa-B: Homeostatic Model Assessment of β-cell function

### Differences in 24h-MB composition between adults with T2DM and controls (aim 1)

The average sleep duration for both groups was approximately 8 hours/night. Adults with T2DM spent on average 11 hours/day on SB, four hours/day LPA, and 30 minutes/day on MVPA **(Table 1)**. A visual representation of the 24h-MBs of both groups can be found in **Supplementary file 2.** Considering the final model controlling for six confounders (sex, age, education, income, smoking status and environment) and one covariate (daylight), a significant difference in the 24h-MB composition between both groups was found (MANOVA test statistic F=3.21, p=0.026; **table 2**). Within their total composition, adults with T2DM spent significant less time in LPA (-34.7 min/day) and MVPA (-24.1 min/day) compared to the control group (**Figure 2.B**). The variation matrix of the 24h-MBs can be found in **Supplementary file 3**.

**Figure 2.**
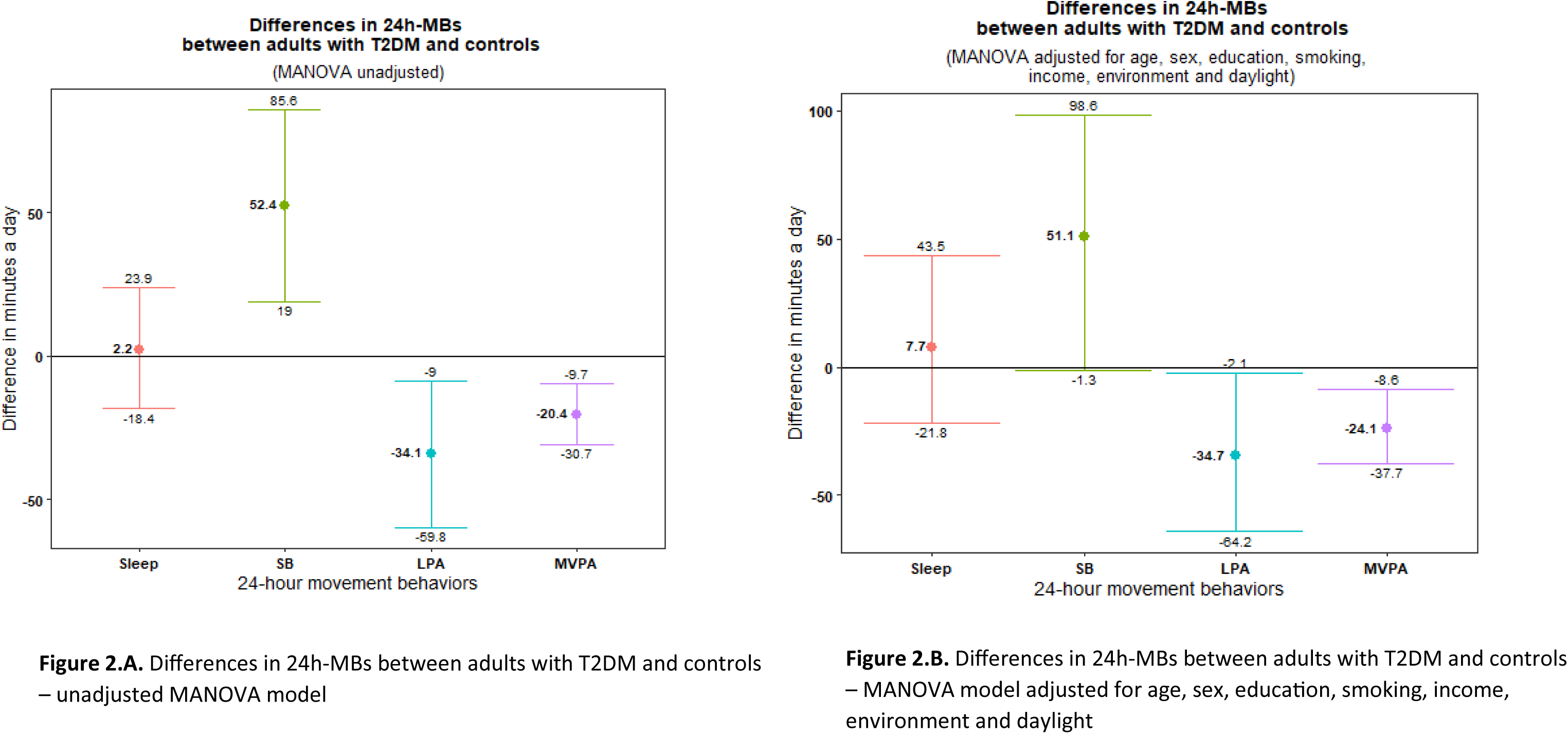
Differences in 24h-MB composition between adults with T2DM and controls. *Abbreviations:* 24h-MB, 24-hour movement behaviors, T2DM, type 2 diabetes mellitus; MANOVA, Multivariate Analysis of Variance; SB, sedentary behavior; LPA, light physical activity; MVPA, moderate to vigorous physical activity

**Table 2.** Differences in 24-hour movement behavior compositions between adults with T2DM and controls.

| Model | F | P-value |
| --- | --- | --- |
| Model 1 | 5.395 | 0.002 |
| Model 2 | 3.207 | 0.026 |

Model 1 Multivariate Analysis of Variance without covariates, Model 2 Multivariate Analysis of Variance adjusted for six confounders (i.e. sex, age, education, smoking status, environment) and one covariate (i.e. mean daylight).

### Associations between cardiometabolic health and 24h-MB features (aim 2)

#### The 24h-MB composition

##### Adults with T2DM

The 24h-MB composition of adults with T2DM was significantly associated with HDL-cholesterol levels (p=0.03) (**Table 3**). **Figure 3.A** illustrates the estimated mean changes in HDL-cholesterol resulting from theoretical reallocations of up to 30 minutes from one behavior to another, keeping the other behaviors constant. Reallocating 30 minutes from any behavior to MVPA is associated with a significant increase in HDL-cholesterol (sleep: 5.05 mg/dl [2.45; 7.80], ES=0.53; SB: 4.53 mg/dl [1.93; 7.27], ES=0.47; LPA: 5.29 mg/dl [2.07; 8.73], ES=0.55).

**Figure 3.**
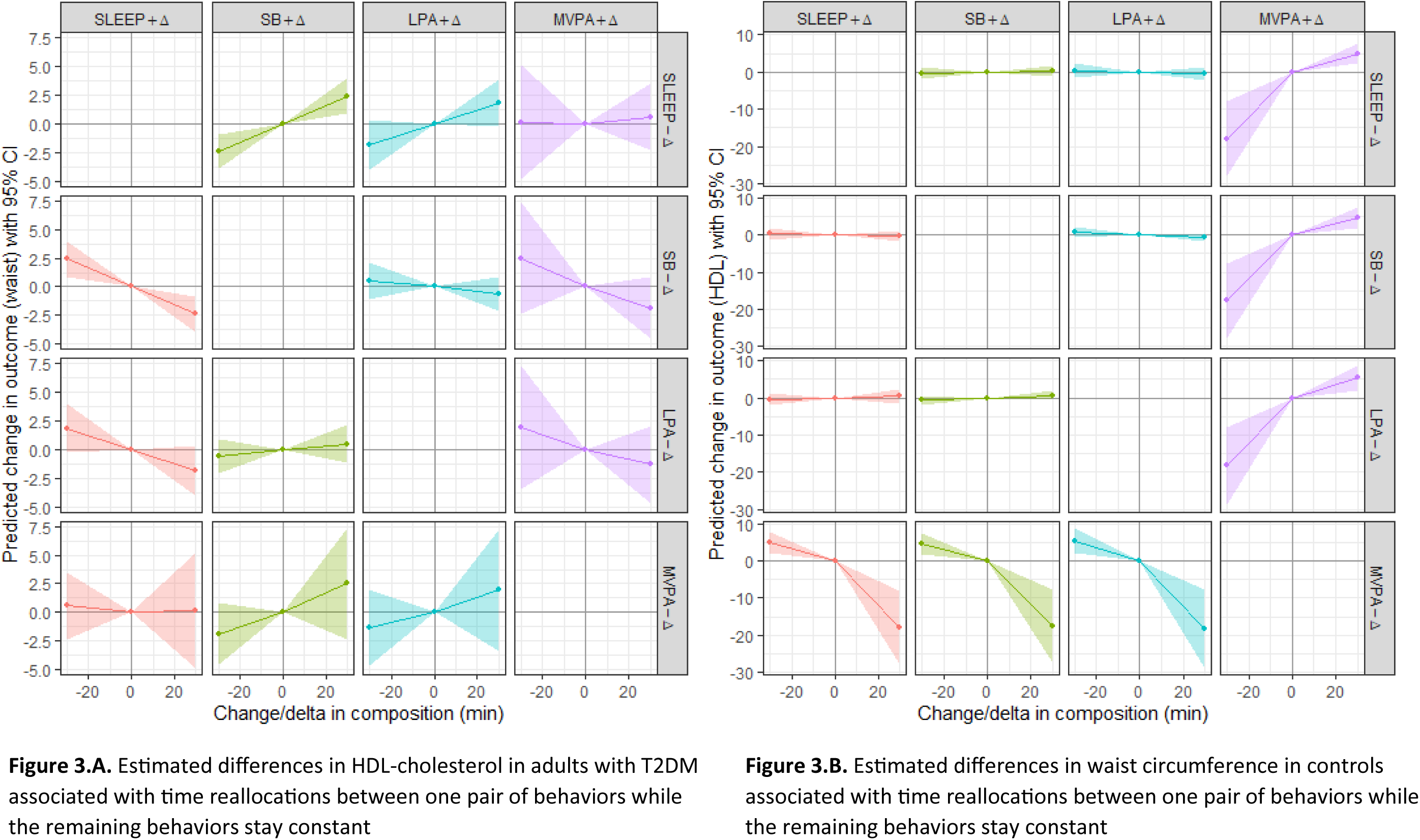
Time reallocation models *Abbreviations:* T2DM, type 2 diabetes mellitus; SB, sedentary behavior; LPA, light physical activity; MVPA, moderate to vigorous physical activity

**Table 3.** Associations between 24h-MB composition and cardiometabolic health parameters at baseline.

| Cardiometabolic health variable | Sum square | F value | p-value | Adjusted p-value<sup>†</sup> |
| --- | --- | --- | --- | --- |
| T2DM (n=52)<sup>‡</sup> |  |  |  |  |
| Body mass index (in kg/m<sup>2</sup>)<sup>⓪</sup> | 0.09 | 1.44 | 0.25 | 0.52 |
| Waist circumference (in cm)<sup>⓪</sup> | 0.04 | 1.10 | 0.36 | 0.52 |
| WHR<sup>⓪</sup> | 0.003 | 0.24 | 0.87 | 0.90 |
| AGE (in arbitrary units) | 1.53 | 1.69 | 0.19 | 0.76 |
| SBP (in mm/Hg) | 162.8 | 0.20 | 0.90 | 0.90 |
| DBP (in mm/Hg) | 339.8 | 0.82 | 0.49 | 0.90 |
| Total cholesterol (in mg/dl)<sup>⓪x</sup> | 0.28 | 1.74 | 0.18 | 0.52 |
| HDL-cholesterol (in mg/dl)<sup>ΦX</sup> | 0.70 | 6.16 | 0.002 | 0.02* |
| LDL-cholesterol (in mg/dl)<sup>ΦX</sup> | 0.18 | 0.59 | 0.63 | 0.69 |
| Triglycerides (in mg/dl)<sup>ΦX</sup> | 0.18 | 0.31 | 0.82 | 0.82 |
| Fasting glucose (in mg/dl)<sup>ΦX</sup> | 0.17 | 1.06 | 0.38 | 0.52 |
| HbA1c (in %)<sup>ΦX</sup> | 0.11 | 1.17 | 0.34 | 0.52 |
| Insulin (in mU/l)<sup>ΦX</sup> | 1.60 | 0.95 | 0.43 | 0.53 |
| HOMA-IR<sup>ΦX</sup> | 1.18 | 1.06 | 0.38 | 0.52 |
| HOMA-B<sup>ΦX</sup> | 1.70 | 2.10 | 0.12 | 0.52 |
| **CONTROL GROUP (n=74)<sup>‡</sup>** |  |  |  |  |
| Body mass index (in kg/m<sup>2</sup>)<sup>Φ</sup> | 0.14 | 2.11 | 0.11 | 0.11 |
| Waist circumference (in cm) | 1773.2 | 3.73 | 0.02 | 0.04 |
| WHR<sup>X</sup> | 0.03 | 1.37 | 0.26 | 0.33 |
| AGE (in arbitrary units) | 0.22 | 2.75 | 0.05 | 0.20 |
| SBP (in mm/Hg) | 1312.6 | 2.13 | 0.11 | 0.22 |
| DBP(in mm/Hg) | 382.2 | 1.18 | 0.33 | 0.33 |

All models were adjusted for sex, age, education, smoking, income, environment and daylight.
<sup>Φ</sup>Log transformed regression model
<sup>‡</sup>Based on the Benjamini-Hochberg procedure
<sup>\*</sup>p<0.05
T2DM: Numerator degrees of freedom: 3, denominator degrees of freedom of F-test: 35 for insulin and HOMA-IR, 37 for LDL-cholesterol and fasting glucose, 38 for BMI, total cholesterol, HDL-cholesterol, triglycerides and HbA1c and 40 for WC and HOMA-B.
Controls: Numerator degrees of freedom: 3, denominator degrees of freedom: 60 for WC and 64 for BMI
<sup>X</sup>*Missings*: see supplementary file 1
*Abbreviations*: 24h-MB, 24-hour movement behavior; T2DM, type 2 diabetes mellitus; HDL, high-density lipoprotein; LDL, low-density lipoprotein; HbA1c, glycated haemoglobin; HOMA-IR, homeostatic model assessment of insulin resistance; HOMA-B, homeostatic model assessment of Beta-cell function

#### Control group

The 24h-MB composition of the controls was significantly associated with WC (p=0.04) (**Table 3**). **Figure 3.B** shows the mean changes in WC resulting from theoretical reallocations of up to 30 minutes.

Reallocating 30 minutes from SB to sleep is associated with a significant decrease in WC (2.42 cm [0.86-3.97], ES=0.34).

#### Average acceleration and intensity gradient

##### Adults with T2DM

Model 3 demonstrated significant associations for either average acceleration or intensity gradient, independent of the alternate metric. This suggests that one of the two metrics may be more strongly related to the specific health outcome being examined. Specifically, intensity gradient was significantly and independently associated with HDL-cholesterol, insulin, HOMA-IR, and HOMA-B, indicating that physical activity intensity, rather than overall volume, was more relevant for these metabolic health markers (**Table 4**).

**Table 4.**
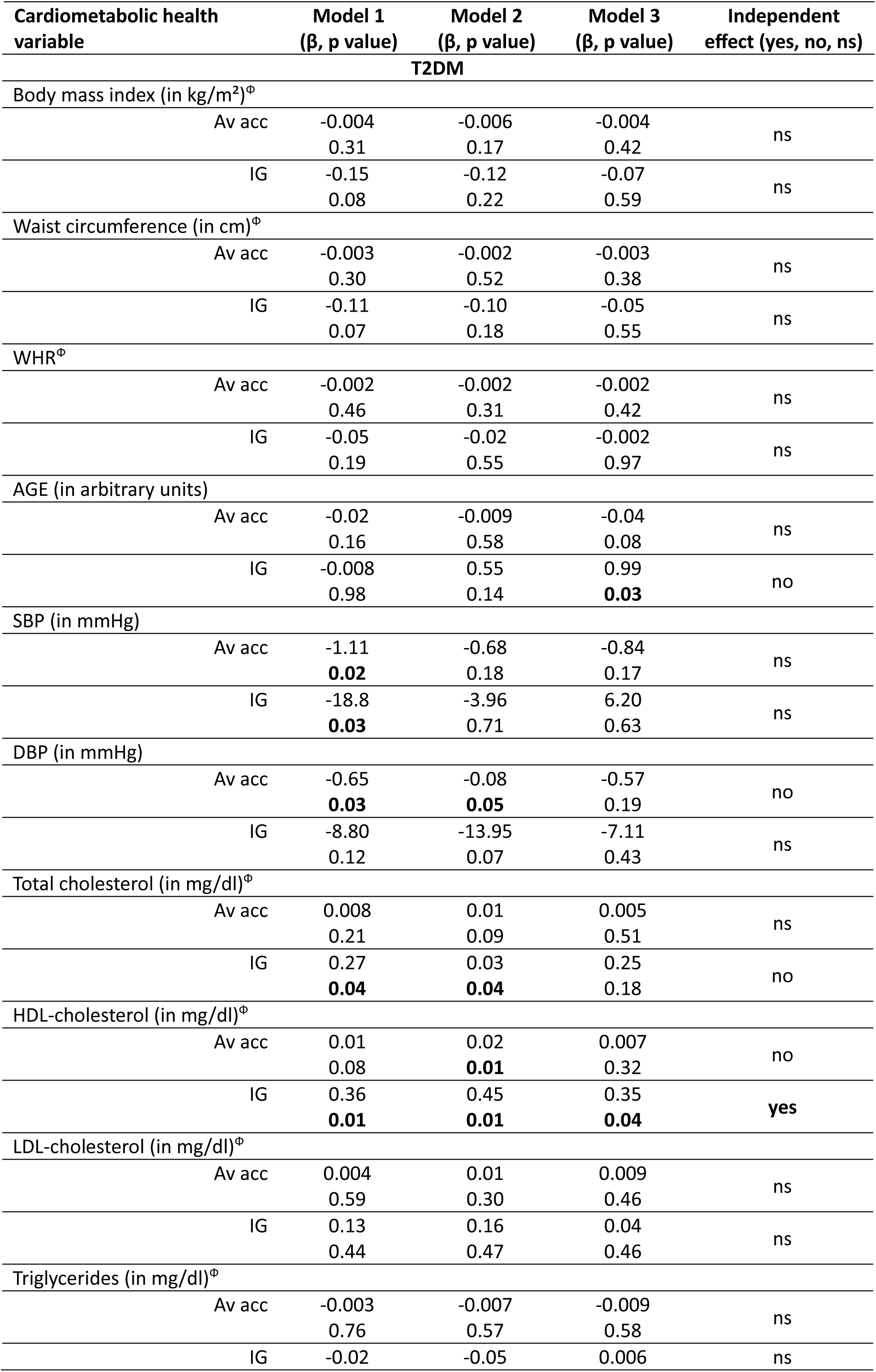
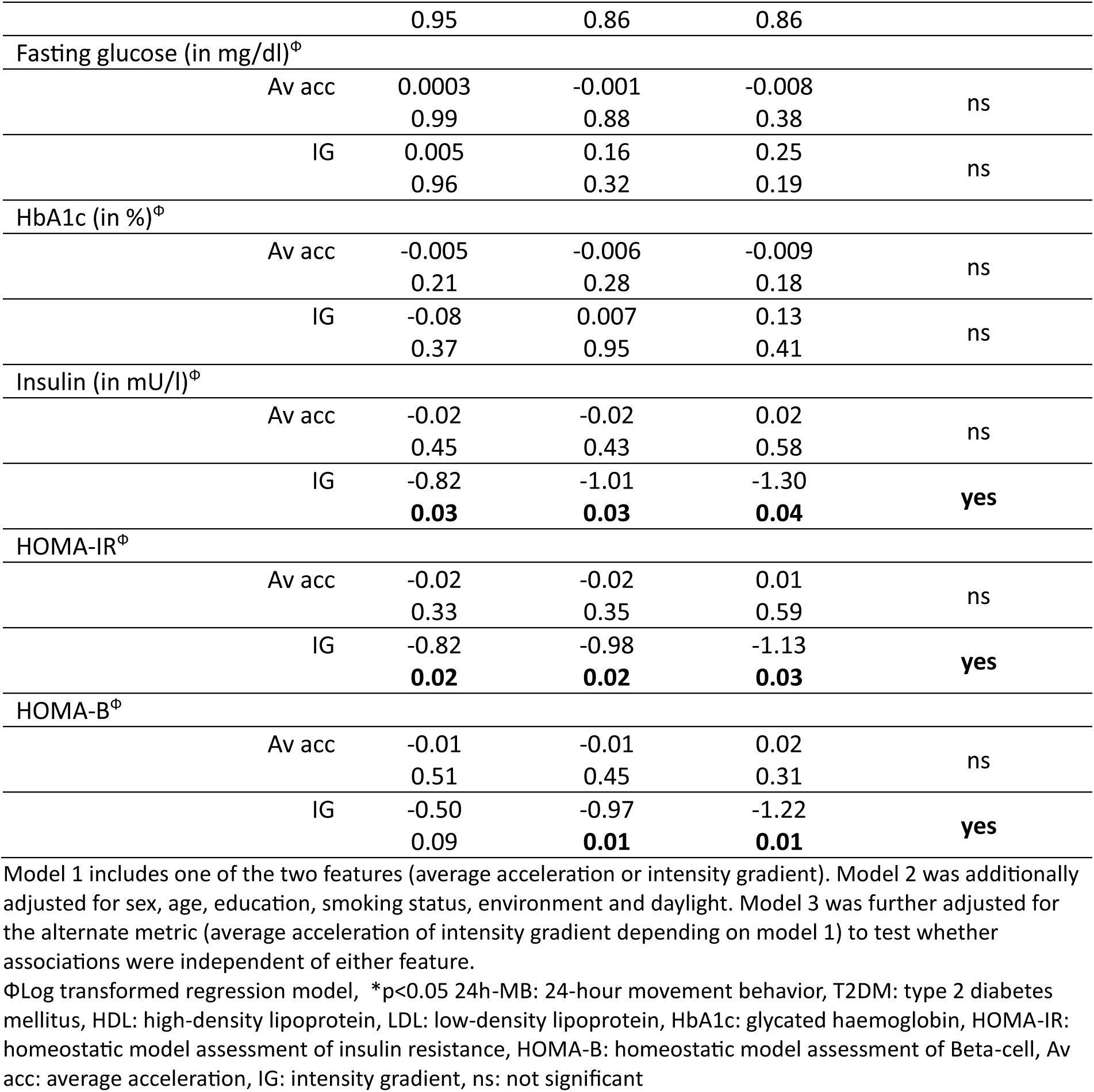
Associations between average acceleration or intensity gradient and cardiometabolic health parameters at baseline in adults with type 2 diabetes mellitus.

| Cardiometabolic health variable | Model 1<br>(β, p value) | Model 2<br>(β, p value) | Model 3<br>(β, p value) | Independent effect (yes, no, ns) |
| --- | --- | --- | --- | --- |

| T2DM |  |  |  |  |
| --- | --- | --- | --- | --- |
| Body mass index (in kg/m<sup>2</sup>)<sup>Φ</sup> |  |  |  |  |
| Av acc | -0.004<br>0.31 | -0.006<br>0.17 | -0.004<br>0.42 | ns |
| IG | -0.15<br>0.08 | -0.12<br>0.22 | -0.07<br>0.59 | ns |
| Waist circumference (in cm)<sup>Φ</sup> |  |  |  |  |
| Av acc | -0.003<br>0.30 | -0.002<br>0.52 | -0.003<br>0.38 | ns |
| IG | -0.11<br>0.07 | -0.10<br>0.18 | -0.05<br>0.55 | ns |
| WHR<sup>Φ</sup> |  |  |  |  |
| Av acc | -0.002<br>0.46 | -0.002<br>0.31 | -0.002<br>0.42 | ns |
| IG | -0.05<br>0.19 | -0.02<br>0.55 | -0.002<br>0.97 | ns |
| AGE (in arbitrary units) |  |  |  |  |
| Av acc | -0.02<br>0.16 | -0.009<br>0.58 | -0.04<br>0.08 | ns |
| IG | -0.008<br>0.98 | 0.55<br>0.14 | 0.99<br>**0.03** | no |
| SBP (in mmHg) |  |  |  |  |
| Av acc | -1.11<br>**0.02** | -0.68<br>0.18 | -0.84<br>0.17 | ns |
| IG | -18.8<br>**0.03** | -3.96<br>0.71 | 6.20<br>0.63 | ns |
| DBP (in mmHg) |  |  |  |  |
| Av acc | -0.65<br>**0.03** | -0.08<br>**0.05** | -0.57<br>0.19 | no |
| IG | -8.80<br>0.12 | -13.95<br>0.07 | -7.11<br>0.43 | ns |
| Total cholesterol (in mg/dl)<sup>Φ</sup> |  |  |  |  |
| Av acc | 0.008<br>0.21 | 0.01<br>0.09 | 0.005<br>0.51 | ns |
| IG | 0.27<br>**0.04** | 0.03<br>**0.04** | 0.25<br>0.18 | no |
| HDL-cholesterol (in mg/dl)<sup>Φ</sup> |  |  |  |  |
| Av acc | 0.01<br>0.08 | 0.02<br>**0.01** | 0.007<br>0.32 | no |
| IG | 0.36<br>**0.01** | 0.45<br>**0.01** | 0.35<br>**0.04** | yes |
| LDL-cholesterol (in mg/dl)<sup>Φ</sup> |  |  |  |  |
| Av acc | 0.004<br>0.59 | 0.01<br>0.30 | 0.009<br>0.46 | ns |
| IG | 0.13<br>0.44 | 0.16<br>0.47 | 0.04<br>0.46 | ns |
| Triglycerides (in mg/dl)<sup>Φ</sup> |  |  |  |  |
| Av acc | -0.003<br>0.76 | -0.007<br>0.57 | -0.009<br>0.58 | ns |
| IG | -0.02 | -0.05 | 0.006 | ns |

|  |  |  |  |  |  |
| --- | --- | --- | --- | --- | --- |
|  |  | 0.95 | 0.86 | 0.86 |  |
| Fasting glucose (in mg/dl)<sup>Φ</sup> |  |  |  |  |  |
|  | Av acc | 0.0003 | -0.001 | -0.008 | ns |
|  |  | 0.99 | 0.88 | 0.38 |  |
|  | IG | 0.005 | 0.16 | 0.25 | ns |
|  |  | 0.96 | 0.32 | 0.19 |  |
| HbA1c (in %)<sup>Φ</sup> |  |  |  |  |  |
|  | Av acc | -0.005 | -0.006 | -0.009 | ns |
|  |  | 0.21 | 0.28 | 0.18 |  |
|  | IG | -0.08 | 0.007 | 0.13 | ns |
|  |  | 0.37 | 0.95 | 0.41 |  |
| Insulin (in mU/l)<sup>Φ</sup> |  |  |  |  |  |
|  | Av acc | -0.02 | -0.02 | 0.02 | ns |
|  |  | 0.45 | 0.43 | 0.58 |  |
|  | IG | -0.82 | -1.01 | -1.30 | yes |
|  |  | 0.03 | 0.03 | 0.04 |  |
| HOMA-IR<sup>Φ</sup> |  |  |  |  |  |
|  | Av acc | -0.02 | -0.02 | 0.01 | ns |
|  |  | 0.33 | 0.35 | 0.59 |  |
|  | IG | -0.82 | -0.98 | -1.13 | yes |
|  |  | 0.02 | 0.02 | 0.03 |  |
| HOMA-B<sup>Φ</sup> |  |  |  |  |  |
|  | Av acc | -0.01 | -0.01 | 0.02 | ns |
|  |  | 0.51 | 0.45 | 0.31 |  |
|  | IG | -0.50 | -0.97 | -1.22 | yes |
|  |  | 0.09 | 0.01 | 0.01 |  |

ΦLog transformed regression model, \*p<0.05 24h-MB: 24-hour movement behavior, T2DM: type 2 diabetes mellitus, HDL: high-density lipoprotein, LDL: low-density lipoprotein, HbA1c: glycated haemoglobin, HOMA-IR: homeostatic model assessment of insulin resistance, HOMA-B: homeostatic model assessment of Beta-cell, Av acc: average acceleration, IG: intensity gradient, ns: not significant

##### Control group

No significant associations were found between any of the cardiometabolic health variables and average acceleration or intensity gradient among the control group (**Table 5**).

**Table 5.** Associations between average acceleration or intensity gradient and cardiometabolic health parameters at baseline in controls.

| CONTROL GROUP |  |  |  |  |  |
| --- | --- | --- | --- | --- | --- |
| Body mass index (in kg/m²)<sup>Φ</sup> |  |  |  |  |  |
|  | Av acc | -0.002 | -0.004 | -0.004 | ns |
|  |  | 0.28 | 0.18 | 0.32 |  |
|  | IG | -0.08 | -0.07 | 0.008 | ns |
|  |  | 0.18 | 0.37 | 0.94 |  |
| Waist circumference (in cm) |  |  |  |  |  |
|  | Av acc | -0.41<br>0.08 | -0.03<br>0.18 | -0.28<br>0.46 | ns |
|  | IG | -13.55<br>0.02 | -7.87<br>0.24 | -2.82<br>0.79 | ns |
| WHR |  |  |  |  |  |
|  | Av acc | -0.001<br>0.23 | -0.001<br>0.48 | -0.001<br>0.59 | ns |
|  | IG | -0.07<br>0.05 | -0.01<br>0.64 | 0.003<br>0.95 |  |
| AGE (in arbitrary units) |  |  |  |  |  |
|  | Av acc | -0.006<br>0.44 | 0.001<br>0.89 | 0.005<br>0.67 | ns |
|  | IG | -0.34<br>0.09 | -0.05<br>0.82 | -0.14<br>0.64 | ns |
| SBP (in mmHg) |  |  |  |  |  |
|  | Av acc | -0.29<br>0.24 | -0.19<br>0.53 | -0.37<br>0.40 | ns |
|  | IG | -5.40<br>0.39 | -0.39<br>0.96 | 6.04<br>0.40 | ns |
| DBP (in mmHg) |  |  |  |  |  |
|  | Av acc | 0.23<br>0.20 | -0.05<br>0.80 | -0.35<br>0.26 | ns |
|  | IG | 7.13<br>0.11 | 3.99<br>0.45 | 10.08<br>0.18 | ns |

ΦLog transformed regression model, \*p<0.05 24h-MB: 24-hour movement behavior, Av acc: average acceleration, IG: intensity gradient, ns: not significant

### Longitudinal changes in 24h-MBs in adults with T2DM (aim 3)

At T1, 37 adults (65.0 SD 9.6 years, 27% female, BMI 29.3 SD 4.6 kg/m²) with T2DM provided valid accelerometer data with 22 participants (67.0 SD 7.7 years, 22.7% female, BMI 28.6 SD 3.9 kg/m²) doing so at T2. No significant changes in composition were found after one (n=37, Wald chi²=0.45, p=0.93) or two years follow-up (n=22, Wald chi²=3.13, p=0.79) (**Table 6**). Drop-out analysis (Supplementary file 4) showed no significant differences in sex, age or body composition between the ones who completed and dropped-out after at T1. At T2, participants who completed were significantly older (p=0.022) compared to the ones who dropped-out (67.09±7.68 vs. 60.33±11.65, resp.).

**Table 6.**
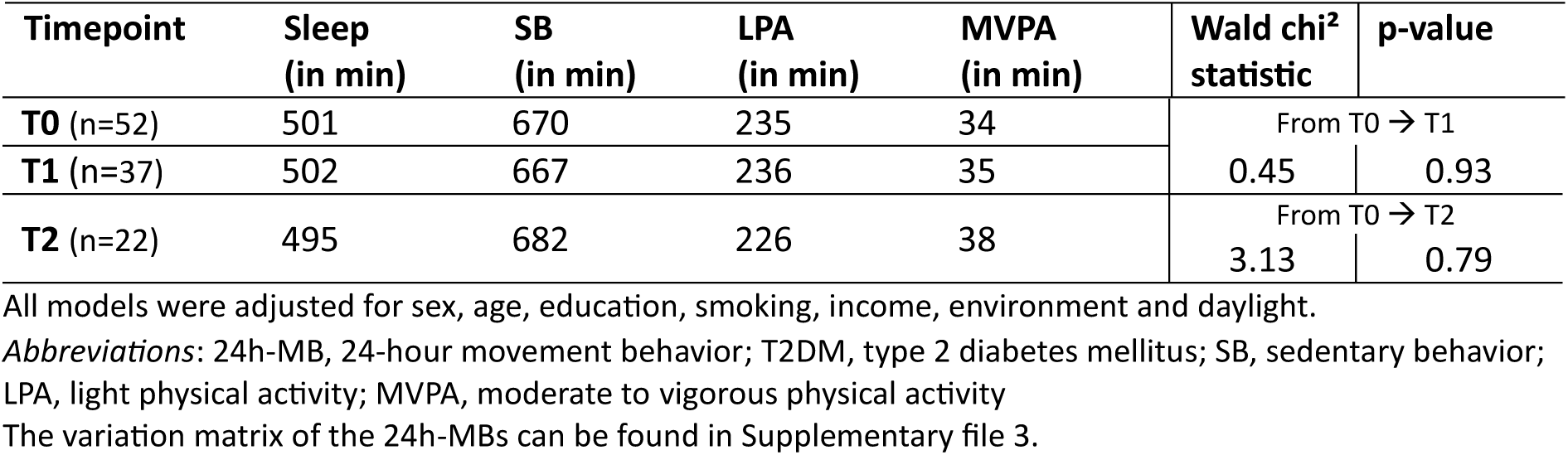
Longitudinal 24h-MB composition in adults with T2DM.

| Timepoint | Sleep<br>(in min) | SB<br>(in min) | LPA<br>(in min) | MVPA<br>(in min) | Wald chi<sup>2</sup><br>statistic | p-value |
| --- | --- | --- | --- | --- | --- | --- |
| **T0** (n=52) | 501 | 670 | 235 | 34 | From T0 → T1 |  |
| **T1** (n=37) | 502 | 667 | 236 | 35 | 0.45 | 0.93 |
| **T2** (n=22) | 495 | 682 | 226 | 38 | From T0 → T2 |  |
|  |  |  |  |  | 3.13 | 0.79 |

All models were adjusted for sex, age, education, smoking, income, environment and daylight.
*Abbreviations:* 24h-MB, 24-hour movement behavior; T2DM, type 2 diabetes mellitus; SB, sedentary behavior; LPA, light physical activity; MVPA, moderate to vigorous physical activity
The variation matrix of the 24h-MBs can be found in Supplementary file 3.

## Discussion

This is the first study directly comparing 24h-MBs between adults with T2DM and adults without diabetes. Our results highlight 24h-MBs differences between both groups with adults with T2DM spending significantly less time in LPA end MVPA compared to controls. These results align with a systematic review by Kennerly and Kirk [19], which demonstrated that adults with T2DM engage less PA (both LPA and MVPA, objectively measured) compared to controls. For example, one included study comparing objectively measured PA between adults with and without T2D, showed that adults with T2DM spend on average 28 min/day less in MVPA (p=0.002) than adults without T2DM, which is comparable with our findings [36]. This difference in PA between adults with and without T2DM can be explained by factors such as physical complications (i.e. neuropathy, weaker muscle strength) and mental challenges (i.e. anxiety) associated with T2DM [37–39]. Moreover, Kennerly and Kirk [19] also reported that adults with T2DM spent more time in SB compared to their adults without diabetes. This aligns with our findings, which show a difference in SB (+51 min/day) between the two groups, although it is not statistically significant. Previous research also showed that adults with T2DM tend to sleep shorter compared to adults without diabetes [40–43]. This may be due to the higher prevalence of sleep disorders such as sleep apnea and insomnia in adults with T2DM [43]. In our study, adults with T2DM had an average sleep duration of 8 hours which was not significantly different from that of the controls. Discrepancy with existing literature may be explained by differences in measurement methods, data analysis methods and sample characteristics.

The 24h-MB composition of adults with T2DM in our study is comparable with the composition of other research among this population, with the exception of MVPA, which is higher (+16-26 min) in our study compared to the other studies [5, 15]. Although there were few differences in the baseline characteristics (e.g. age and sex distribution) between the above studies and our study, differences in results may be due to variations in data analysis methods of movement behaviors (Väha-ÿpa versus Freedson [5] or Troiano [15] cut-points), which has been shown to have big implications on the average time spent in 24h-MBs [44]. Moreover differences in local habits (e.g. cycling in the Netherlands [15], hiking in Sweden (hiking [5])) and sample size (52 vs 175 [5] vs 1001 [15]) can contribute to differences in results.

Our study found a significant association between HDL-cholesterol and the 24h-MB composition in adults with T2D. Significant increases in HDL-cholesterol where found when reallocating 30 minutes from sleep, SB or LPA into MVPA. This result aligns with the fact that HDL-cholesterol is more affected by lifestyle changes (compared to medication) and the well-established linear dose-response relationship between PA and HDL cholesterol [45, 46]. The observed increase of 4.53-5.29 mg/dl in HDL cholesterols is clinically relevant as even small increase in HDL cholesterol are associated with a decrease in the risk of cardiovascular events [47]. Our results are in line with the findings of the study of Willems, Verbestel [15] where significant increases in HDL-cholesterol were observed when reallocating time from SB or sleep into LPA or MVPA. However, the magnitude of these changes where notably smaller compared to our results. Differences in sample size and data analysis approaches may explain the observed variation in associations and time reallocation outcomes. Nevertheless, our study and others consistently show that reallocating time from any other behavior to MVPA has the strongest effects on HDL cholesterol and overall health outcomes, as confirmed by the review of Miatke, Olds [48]. Moreover we found an independent positive association between intensity gradient and HDL-cholesterol, and a negative independent associations with insulin, HOMA-IR and HOMA-B. This suggests that PA intensity rather than PA volume plays a more significant role in influencing these variables. This reflects the potential beneficial effects of intensity on the lipid profile and insulin sensitivity which is already shown in experimental research. While both diet and PA contribute to improvement in lipid profiles, HDL-cholesterol is known to be more responsive to exercise than to dietary changes [49, 50]. Regarding insulin sensitivity, both intensity and volume are important although higher intensities appear to have a greater impact [51]. However it is also important to consider other behaviors (e.g. LPA) in improving health, as highlighted in the consensus report from the American Diabetes Association and European Association for the Study of Diabetes [9]. Although our study did not demonstrate this, the study mentioned above by Willems, Verbestel [15] suggested that reallocating time from SB to LPA can already yield significant health benefits (i.e. lower BMI, WC and triglycerides). These changes may be more achievable for people with reduced exercise capacity (i.e. adults with T2D), compared to increasing MVPA [15].

Significant associations were found between the 24h-MB composition and WC in healthy controls. Similar to results of other studies, theoretical reallocations from SB to sleep were associated with reductions in WC, which is associated with a reduction in the risk of T2DM [52, 53]. Despite these results, the reallocation from other behaviors to sleep, or vice versa, should be done with caution. The relationship between sleep and health outcomes is U-shaped, meaning that both long (mostly defined as ≥9h/night) and short (mostly defined as <7h/night) sleep durations are associated with negative health outcomes (e.g. cardiovascular disease, obesity) [54, 55]. This has also been demonstrated in adults with T2DM [15]. In this study, we did not make a distinction between long and short sleepers due to the small sample size and limited variation in sleep duration (490 minutes [range 444-510]).

No changes in 24h-MB after two years of follow up were observed. These results align with a longitudinal study that examined PA patterns among individuals with prediabetes and T2D, where the majority of participants (78%, n=131) maintained their PA levels over the two-year follow-up period [56]. In contrast, a Finnish study examining 24h-MBs in adults transitioning from work to retirement reported a decrease in LPA and MVPA along with an increase in SB and sleep after the transition to retirement [57]. Since most participants in our study were already retired, their consistent lifestyle patterns may explain the minimal changes in 24h-MBs observed after two years of follow up. Furthermore, no significant changes were observed in cardiometabolic and adiposity variables after two years of follow-up, which may indicate a stable management (i.e. 24h-MBs) of their T2D. However, there remains considerable room for improvement in this population, as their 24h-MB composition is still not optimal (e.g. ±11 hours/day SB).

This is the first study comparing objectively collected 24h-MBs between adults with T2DM and controls. Objectively collected raw data (Actigraph GT3X+) was processed by a commonly used R package GGIR [23]. By not using subjective measurement methods, except the sleep diary, the risk of recall- and social desirability bias was limited. Another strength of this study was the availability of longitudinal data which provided valuable insights into behavioral changes over time. This study also has some limitations. An a priori sample size calculation revealed a minimum sample size of 92 participants for the longitudinal follow-up study (α= 0.05, β=0.80, Mean Difference PA= 627; Standard Deviation= 2116; correlation=0.5)[58]. However, only a small sample of adults with T2DM was recruited at baseline and participated in the follow-up measurements due to recruitment issues. In addition to that, there was a high drop-out rate at both the one-year (29%) and two-year follow-up (40%) which resulted in small sample sizes. These small sample size may have resulted in type II errors due to low statistical power. Furthermore, the sampling method may have induced motivation bias. Additionally, the average HbA1c was good and the majority of participants (71.1%) were only taking one oral antidiabetic medication which suggests a well-controlled T2DM population and may contribute to finding few associations. Furthermore, the participants could choose whether the data collection took place at the Ghent University Hospital or at the participant’s home. The location could potentially influence certain measurements, such as BP. Another limitation is absence of follow-up data and blood samples in the control group. Since no blood samples were taken in the control group, it is possible that these individuals have prediabetes. Additionally, we cannot say whether the associations found between the 24h-MBs and the blood parameters in the adults with T2DM are due to their diabetes or not. Future research should focus on measuring 24h-MBs in larger groups of adults with T2DM over an extended follow-up period, as this will give insight into how the behavior of individuals changes as their condition progresses, which may be valuable for the T2DM management and intervention development.

## Conclusion

There is a significant difference in 24h-MBs between adults with and without T2DM. The adults with T2DM showed less favorable 24h-MBs, which remained stable over a two years underscoring the need for tailored 24h-MB guidelines for this population. HDL-cholesterol and WC were associated with the 24h-MBs of adults with T2DM and controls, respectively. Reallocating time from any other behavior to MVPA was associated with significant improvements in HDL-cholesterol in the adults with T2DM while significant reductions in WC of control adults were found when shifting time from SB to sleep. Although only a few associations were found, our data add to the existing literature to underscore the need to prioritize a healthy 24h-MBs in adults with T2DM.

## Supporting information

Supplementary file 1. Missing_data_baseline_characteristics

Supplementary file 2. 24h-MBs in adults with T2DM and controls

Supplementary file 3. Variation matrix

Supplementary file 4. Drop-out analysis

## Data Availability

All data produced in the present study are available upon reasonable request to the authors

## Author contributions

L. Bogaert, M. De Craemer and I. Willems conceived the initial idea for this paper and designed the protocol L. Bogaert and I. Willems collected the data. L. Bogaert and I. Willems analyzed the data. L. Bogaert, M. De Craemer and I. Willems wrote the manuscript. All authors reviewed and approved the final version of the manuscript.

## Acknowledgements

Not applicable

