## Supplementary file 1. Missing_data_baseline_characteristics for "24-hour movement behaviors and cardiometabolic health in adults with type 2 diabetes: a comparative cross-sectional and longitudinal analysis"

**Supplementary file 1.** Missing data of baseline characteristics.

| Variable | T2DM (n=52) | Control (n=74) |
|---|---|---|
| Environment (n) | 1 | |
| Education (n) | 1 | |
| Work status (n) | 1 | |
| Household income (n) | 1 | 2 |
| Smoking status (n) | 1 | |
| Cardiometabolic health | | |
| Waist to hip ratio (n) | | 2 |
| HbA1c (n) | 1 | |
| Insulin (n) | 4 | |
| Fasting glucose (n) | 2 | |
| Total cholesterol (n) | 1 | |
| HDL-cholesterol (n) | 1 | |
| LDL-cholesterol (n) | 2 | |
| Triglycerides (n) | 1 | |
| HOMA-IR (n) | 4 | |
| HOMA-B (n) | 4 | |
| Medication | | |
| Medication intake (yes/no) (n) | | 2 |
| Lipid lowering medication (n) | 1 | 2 |
| Blood pressure lowering medication (n) | 1 | 2 |
| Anticoagulant medication (n) | 1 | 2 |

*Abbreviations*: T2D: type 2 diabetes; LDL-cholesterol: low-density lipoprotein cholesterol; HDL-cholesterol: high-density lipoprotein cholesterol; HOMA-IR: Homeostatic Model Assessment of Insulin Resistance; Homa-B: Homeostatic Model Assessment of β-cell function
