## Supplementary file 2. 24h-MBs in adults with T2DM and controls for "24-hour movement behaviors and cardiometabolic health in adults with type 2 diabetes: a comparative cross-sectional and longitudinal analysis"

**Supplementary 2.** 24-hour movement behaviors in adults with T2DM and controls.

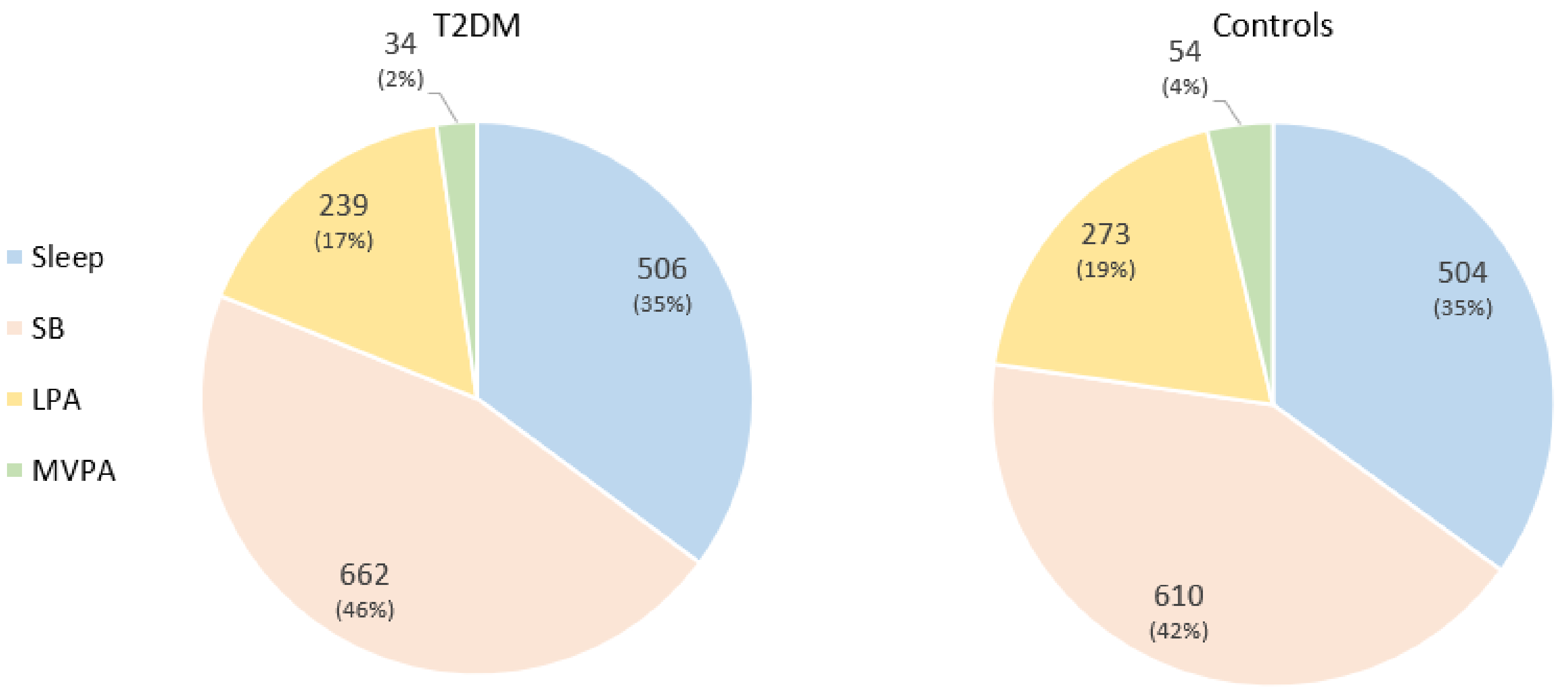


Data is represented as mean minutes/day (percentage)

*Abbreviations:* T2DM, type 2 diabetes mellitus; SB, sedentary behavior; LPA, light physical activity; MVPA, moderate to vigorous physical activity
