## Supplementary file 3. Variation matrix for "24-hour movement behaviors and cardiometabolic health in adults with type 2 diabetes: a comparative cross-sectional and longitudinal analysis"

**Supplementary file 3.** Variation matrices

Variation matrix representing the co-dependency between 24h-MBs at baseline

| | **Sleep** | **SB** | **LPA** | **MVPA** |
|---|---|---|---|---|
| **Total sample at baseline** | | | | |
| Sleep | 0.000 | 0.053 | 0.114 | 0.598 |
| SB | 0.053 | 0.000 | 0.169 | 0.686 |
| LPA | 0.114 | 0.169 | 0.000 | 0.459 |
| MVPA | 0.598 | 0.686 | 0.459 | 0.000 |
| **Adults with T2DM at baseline** | | | | |
| Sleep | 0.000 | 0.048 | 0.143 | 0.696 |
| SB | 0.048 | 0.000 | 0.180 | 0.736 |
| LPA | 0.143 | 0.180 | 0.000 | 0.574 |
| MVPA | 0.696 | 0.736 | 0.574 | 0.000 |
| **Healthy controls at baseline** | | | | |
| Sleep | 0.000 | 0.054 | 0.087 | 0.443 |
| SB | 0.054 | 0.000 | 0.145 | 0.531 |
| LPA | 0.087 | 0.145 | 0.000 | 0.336 |
| MVPA | 0.443 | 0.531 | 0.336 | 0.000 |

Variation matrix representing the co-dependency between 24h-MBs at follow-up

| | **Sleep** | **SB** | **LPA** | **MVPA** |
|---|---|---|---|---|
| **Adults with T2DM – timepoint 1** | | | | |
| Sleep | 0.000 | 0.033 | 0.059 | 0.467 |
| SB | 0.033 | 0.000 | 0.019 | 0.346 |
| LPA | 0.059 | 0.019 | 0.000 | 0.398 |
| MVPA | 0.467 | 0.346 | 0.398 | 0.000 |
| **Adults with T2DM – timepoint 2** | | | | |
| Sleep | 0.000 | 0.042 | 0.050 | 0.263 |
| SB | 0.042 | 0.000 | 0.015 | 0.194 |
| LPA | 0.050 | 0.015 | 0.000 | 0.302 |
| MVPA | 0.263 | 0.194 | 0.302 | 0.000 |

*Abbreviations:* 24h-MBs, 24-hour movement behaviors; SB, sedentary behavior; LPA, light physical activity; MVPA, moderate to vigorous physical activity; T2DM, type 2 diabetes mellitus
