## Supplementary file 4. Drop-out analysis for "24-hour movement behaviors and cardiometabolic health in adults with type 2 diabetes: a comparative cross-sectional and longitudinal analysis"

**Supplementary file 4.** Drop-out analysis in adults with T2DM

| FROM BASELINE TO FOLLOW-UP 1 | | | |
|---|---|---|---|
| **Variable** | **Completed (n=37)** | **Dropped out (n=15)** | **P-value** |
| BMI | 29.69 (4.70) | 31.65 (6.84) | 0.242 |
| BMI categories (%) | | | 0.343 |
| - Normal weight (<25kg/m²) | 7 (18.9) | 2 (13.3) | |
| - Overweight (25-29.9 kg/m²) | 16 (43.2) | 4 (26.7) | |
| - Obese (≥30 kg/m²) | 14 (37.8) | 9 (60.0) | |
| Waist circumference (in cm) | 105.73 (11.00) | 111.79 (18.39) | 0.148 |
| Waist to hip ratio | 0.99 (0.07) | 0.98 (0.10) | 0.747 |
| Systolic blood pressure | 141.31 (14.02) | 128.40 (17.23) | 0.007* |
| Diastolic blood pressure | 83.58 (8.76) | 81.23 (12.12) | 0.438 |
| Advanced Glycation Endproducts | 2.75 (0.64) | 2.57 (0.48) | 0.333 |
| HbA1c (mmol/mol) | 52.58 (6.88) | 50.21 (12.62) | 0.398 |
| Insulin | 14.41 (11.21) | 26.29 (19.56) | 0.013 |
| Fasting glucose (mg/dl) | 131.19 (26.88) | 122.46 (43.70) | 0.404 |
| Total cholesterol (mg/dl) | 148.58 (43.21) | 158.21 (27.95) | 0.445 |
| HDL- cholesterol (mg/dl) | 48.17 (13.21) | 46.00 (10.43) | 0.585 |
| LDL-cholesterol (mg/dl) | 81.00 (32.77) | 88.41 (20.42) | 0.437 |
| Triglycerides (mg/dl) | 117.44 (56.57) | 150.14 (111.11) | 0.175 |
| HOMA-IR | 2.10 (1.51) | 2.37 (1.56) | 0.591 |
| HOMA-B | 76.34 (44.02) | 106.20 (39.63) | 0.044* |
| T2DM duration (in years) | 11.97 (8.86) | 8.73 (5.92) | 0.200 |
| Age | 64.62 (9.64) | 59.73 (12.30) | 0.133 |
| Sex | | | 0.557 |
| - Men | 27 (73.0) | 9 (60.0) | |
| - Women | 10 (27.0) | 6 ( 40.0) | |
| Environment | | | 0.639 |
| - City | 14 (37.8) | 8 (50.0) | |
| - Rural area | 23 (62.2) | 7 (50.0) | |
| Education | | | 0.700 |
| - Low | 19 (51.4) | 9 ( 64.3) | |
| - High | 18 (48.6) | 6 (35.7) | |
| Work status | | | 1.000 |
| - Working | 23 (62.2) | 10 (64.3) | |
| - Retired or unemployed | 14 (37.8) | 5 (35.7) | |
| Household income | | | 0.551 |
| - Low | 6 (16.2) | 4 (28.6) | |
| - High | 31 (83.8) | 11 (71.4) | |
| Smoking status | | | 0.520 |
| - Ex-smoker | 12 (32.4) | 5 ( 35.7) | |
| - Smoker | 2 ( 5.4) | 2 ( 14.3) | |
| - Non-smoker | 23 (62.2) | 7 ( 50.0) | |
| Medication (yes) | 37 (100) | 15 (100) | NA |
| Glucose lowering (yes) | 34 (91.9) | 11 (73.3) | 0.184 |
| Lipid lowering (yes) | 23 (62.2) | 9 (60.0) | 1.000 |
| Blood pressure lowering (yes) | 23 (62.2) | 4 (26.7) | 1.000 |
| Anticoagluantia (yes) | 17 (45.9) | 4 (26.7) | 0.331 |
| **FROM BASELINE TO FOLLOW-UP 2** | | | |

| Variable | Completed (n=22) | Dropped out (n=30) | P-value |
|---|---|---|---|
| BMI | 28.59 (3.99) | 31.46 (6.04) | 0.058 |
| BMI categories (%) | | | 0.292 |
| Normal weight (<25kg/m²) | 5 (22.7) | 4 ( 13.3) | |
| Overweight (25-29.9 kg/m²) | 10 (45.5) | 10 ( 33.3) | |
| Obese (≥30 kg/m²) | 7 (31.8) | 16 ( 53.3) | |
| Waist circumference (in cm) | 103.24 (11.12) | 110.47 (14.70) | 0.059 |
| Waist to hip ratio | 0.99 (0.08) | 0.99 (0.08) | 0.986 |
| Systolic blood pressure | 139.39 (14.91) | 133.97 (19.53) | 0.282 |
| Diastolic blood pressure | 82.55 (8.99) | 81.98 (12.62) | 0.859 |
| Advanced Glycation Endproducts | 2.80 (0.68) | 2.62 (0.52) | 0.300 |
| HbA1c (mmol/mol) | 51.32 (5.96) | 51.93 (10.69) | 0.810 |
| Insulin | 9.75 (5.14) | 23.97 (16.47) | <0.001* |
| Fasting glucose (mg/dl) | 128.36 (29.06) | 129.00 (34.02) | 0.945 |
| Total cholesterol (mg/dl) | 159.50 (47.15) | 145.45 (31.30) | 0.207 |
| HDL- cholesterol (mg/dl) | 52.59 (14.08) | 43.52 (9.30) | 0.008* |
| LDL-cholesterol (mg/dl) | 86.64 (38.44) | 81.10 (20.63) | 0.517 |
| Triglycerides (mg/dl) | 108.91 (51.44) | 138.72 (88.23) | 0.164 |
| HOMA-IR | 1.42 (0.70) | 2.77 (1.68) | 0.001* |
| HOMA-B | 62.49 (32.35) | 101.12 (45.04) | 0.002* |
| T2DM duration (in years) | 12.27 (6.78) | 10.13 (9.10) | 0.358 |
| Age | 67.09 (7.68) | 60.33 (11.65) | 0.022* |
| Sex | | | 0.440 |
| Men | 17 (77.3) | 19 (63.3) | |
| Women | 5 (22.7) | 11 ( 36.7) | |
| Environment | | | 0.800 |
| City | 10 (45.5) | 12 (37.9) | |
| Rural area | 12 (54.5) | 18 ( 62.1) | |
| Education | | | 0.061 |
| Low | 8 (36.4) | 20 (69.0) | |
| High | 24 (63.6) | 9 (31.0) | |
| Work status | | | 0.180 |
| Working | 5 (22.7) | 13 (44.8) | |
| Retired or unemployed | 17 (77.3) | 17 (55.2) | |
| Household income | | | 1.000 |
| Low | 4 (18.2) | 6 (20.7) | |
| High | 18 (81.8) | 24 (79.3) | |
| Smoking status | | | 0.211 |
| Ex-smoker | 5 (22.7) | 12 ( 41.4) | |
| Smoker | 1 ( 4.5) | 3 ( 10.3) | |
| Non-smoker | 16 (72.7) | 14 ( 48.3) | |
| Medication (yes) | 22 (100) | 30 (100) | NA |
| Glucose lowering (yes) | 19 (86.4) | 26 (86.7) | 1.000 |
| Lipid lowering (yes) | 13 (59.1) | 19 (63.3) | 0.982 |
| Blood pressure lowering (yes) | 15 (68.2) | 18 (60.0) | 0.754 |
| Anticoagluantia (yes) | 13 (59.1) | 8 (26.7) | 0.039* |

*p<0.05, continuous variables were represented as mean (standard deviation), categorical variables as number (percentage). *Abbreviations*: T2DM: type 2 diabetes mellitus; BMI: body mass index; LDL-cholesterol: low-density lipoprotein cholesterol; HDL-cholesterol: high-density lipoprotein cholesterol; HOMA-IR: Homeostatic Model Assessment of Insulin Resistance; Homa-B: Homeostatic Model Assessment of β-cell function
